# The Understanding persistent Pain Where it ResiDes study of low back pain cohort profile

**DOI:** 10.1101/2021.11.24.21266794

**Authors:** Luke C Jenkins, Wei-Ju Chang, Valentina Buscemi, Matthew Liston, Michael Nicholas, Thomas Graven-Nielsen, Paul W Hodges, Valerie C Wasinger, Laura S Stone, Susan G Dorsey, James H McAuley, Siobhan M Schabrun

## Abstract

**Background:** Despite chronic low back pain (LBP) being considered a biopsychosocial condition for diagnosis and management, few studies have investigated neurophysiological or neurobiological risk factors thought to underpin the transition from acute to chronic LBP. The aim of this cohort profile is to describe the methodology, compare baseline characteristics between acute LBP participants and pain-free controls, and compare LBP participants with or without completed follow-up.

**Methods:** 120 individuals experiencing acute LBP and 57 pain-free controls were recruited to participate in the Understanding Persistent Pain Where it Resides (UPWaRD) study. Screening was conducted via email and phone. Neurobiological, psychological, and sociodemographic data were collected at baseline, three- and six-months. LBP status was assessed using the numerical rating scale and Roland-Morris disability questionnaire at three and six-month follow-up.

**Results:** 95 participants (79%) provided outcome data at three-month follow-up and 96 participants (80%) at six-months. Participants who did not complete follow-up at three- and six-months within the UPWaRD LBP cohort had higher psychological distress, higher pain interference, higher levels of moderate physical activity, and reported occupational difficulties due to pain (*P* = <0.05). Compared to controls, LBP participants in the UPWaRD cohort were older, had a higher BMI, a higher prevalence of comorbidities and higher medication usage. Higher depression, anxiety and stress, lower pain self-efficacy and higher pain catastrophizing during acute LBP were correlated with higher six-month pain and disability (*P* = < 0.01).

**Conclusions:** This cohort profile reports baseline characteristics of the UPWaRD LBP and pain-free control cohort.

## 1. INTRODUCTION

The worldwide one-month prevalence of low back pain (LBP) is approximately 23% and 83% of the world’s population will experience LBP at least once during their lifetime [19;33]. Of those who experience an acute episode of LBP, up to 40% will develop chronic or recurrent symptoms [17]. LBP is a leading cause of disability worldwide [58] and associated with substantial economic burden, with $135 billion spent on low back and neck pain in the US in 2017 [9]. Despite the scale of the problem, identifying those with acute LBP who are at risk of chronic or recurrent symptoms remains challenging.

Most cases of LBP have no identifiable pathoanatomical cause or clear nociceptive source that could explain persistent symptoms [32]. This has led to a focus on the identification of psychological, social and demographic risk factors to explain the transition to chronic LBP [2]. Using this approach, the strongest predictors of chronic LBP are high pain severity during the acute-stage [3;10;13;49;59] and a prior history of LBP [53]. Other factors that increase the risk of experiencing an episode of LBP include psychological distress, smoking and physical inactivity [41]. However, these risk factors explain only a small proportion of the variance in LBP outcome [15;24;28;47].

Investigation of biological risk factors in the development of chronic pain has been limited. Although some data are beginning to show systemic inflammation and pain sensitivity interact with psychological features [25;26], the role of sensorimotor neurobiology has not been investigated. The Understanding persistent Pain Where it ResiDes study (UPWaRD study) aimed to recruit and follow a cohort of adults living in Australia who experienced an acute episode of LBP. The primary aim as reported ‘a priori’ in the study protocol [21] was to use this cohort to identify biological (with an emphasis on neurophysiological factors), psychological and sociodemographic risk factors, and/or interactions between these factors during an acute episode of LBP that predict the development of chronic LBP. The neurobiological risk factors selected for investigation in the protocol were those with a putative link to the development of aberrant cortical and spinal neuroplasticity, hypothesized to explain why some individuals develop chronic pain after an acute episode.

Understanding the baseline characteristics of participants included in the UPWaRD trial is important for accurate and transparent reporting of future longitudinal analyses from this cohort. Therefore, the aims of this cohort profile paper were to: i) describe the design, participant recruitment and measurement procedures of the UPWaRD study, including additional measures and data collected on a group of pain-free control participants, ii) to identify which baseline characteristics (health, sociodemographic, psychological and lifestyle factors) differ between individuals with acute LBP and pain-free controls, (iii) to determine whether any baseline features differ between participants who did, and did not, complete 6-month follow-up, and iv) describe the recovery trajectories (pain and disability) of individuals with acute LBP over a period of 6 months.

## 2. METHODOLOGY

### 2.1. Study design

The UPWaRD study was a multicentre, prospective, longitudinal, cohort trial of people with acute (within 6-weeks of pain onset) LBP, and pain-free controls, with three- and six-month follow-up. The study received funding from the National Health and Medical Research Council of Australia (Grant ID: 1059116). All study procedures were approved by the Human Research Ethics Committees of Western Sydney University (H10465) and Neuroscience Research Australia (SSA: 16/002) and in accordance with the Helsinki Declaration of 1975, as revised in 1983. All participants provided written, informed consent for participation in the study and its related procedures.

### 2.2. Recruitment and follow-up

Participants were recruited through flyers around university campuses and the local community, social media posts, local hospitals in South Eastern Sydney and South Western Sydney Local Health Districts, New South Wales, Australia, primary care practitioners (e.g. general practitioners and physiotherapists) and newspaper advertisements. Screening was conducted via email and phone. Potential participants who contacted the research team or were referred from a practitioner were contacted over the phone within 24 hours to discuss the study purpose and methodology. Participants were then sent a detailed participant information sheet and screening form via email. Participants who returned the screening form were considered “screened” and any reason for exclusion was documented.

Acute LBP participants were eligible if they experienced pain in the region of the lower back, superiorly bound by the thoracolumbar junction and inferiorly by the gluteal fold [35]. Pain must have been present for more than 24 hours and persisted for less than six weeks following a period of at least one-month pain-free [8;35;48;61]. All participants with pain referred beyond the inferior gluteal fold underwent a physical examination by a trained physiotherapist (study staff) to identify any sensory or motor deficit of the lower extremity. Participants with suspected lumbosacral radiculopathy characterised by the presence of weakness, loss of sensation, or loss of reflexes associated with a particular nerve root, or a combination of these, were excluded [29]. Individuals who presented with suspected serious spine pathology (e.g. fracture, tumour, cauda equina syndrome), other major diseases/disorders (e.g. schizophrenia, chronic renal disorder, multiple sclerosis), a history of spine surgery, any other chronic pain conditions or contraindications to the use of transcranial magnetic stimulation (TMS) (as described by Keel et al. [22]) were excluded. Control participants were eligible for study inclusion if they met the relevant exclusion criteria above and had not experienced LBP in the past 12 months.

### 2.3. Data collection

Participants completed a laboratory testing session and a battery of questionnaires (online or in person) at baseline, three- and six-months. All variables were measured in a standardised order for all participants and four assessors performed all laboratory sessions between Western Sydney University, Campbelltown Campus or Neuroscience Research Australia. Duration of assessment of all variables was approximately 2.5 hours. Measures were collected within the domains of health (e.g. weight), sociodemographic (e.g. cultural diversity), psychological (e.g. depression, catastrophising, self-efficacy), clinical (Keele StarT Back Screening Tool), neurobiological (e.g. electroencephalography), biological (serum biomarkers), pain processing (e.g. pressure pain sensitivity) and lifestyle (e.g. physical activity - International Physical Activity Questionnaire). Detailed description of all measures obtained in the UPWaRD Cohort and their methodology is described in supplementary material **(Supplemental File: Table S1)**. This Table includes details of which measures were added after registration/protocol publication. Pain-free controls were followed up at three-and six-months to allow comparison of neurobiological and psychological variables between participants with and without LBP, and allow assessment of measurement stability across baseline, three- and six-months in pain-free individuals [7].

In brief, neurobiological measures were selected based upon a theoretical association between cortical and spinal plasticity and the development of chronic LBP and supporting evidence from cross-sectional studies [3;12;16;30;45;55]. For psychological measures, three questionnaires were used to assess specific aspects of psychological status with evidence of relevance to the development of chronic LBP: the 21-Item Depression, Anxiety and Stress Scales Questionnaire (DASS-21) [1;40], the 13-item Pain Catastrophising Scale (PCS) [51], and the 10-item Pain Self-Efficacy Questionnaire (PSEQ) [36]. A commonly used clinical prediction tool, The Keele StarT Back Screening Tool (SBT) was also administered amongst LBP participants at baseline assessment [18]. Sociodemographic, environmental and lifestyle factors were selected based on the Australasian Electronic Persistent Pain Outcomes Collaboration minimum dataset recommendations [52]. Guidelines for that minimum dataset were first developed in 2011 by an expert team, consisting of members of the Faculty of Pain Medicine of the Australian and New Zealand College of Anaesthetists, Australian Pain Society and New Zealand Pain Society. Participants were free to seek and utilise any treatment, and data were collected on healthcare utilization and medication consumption.

Average pain intensity over the week preceding baseline and follow-up assessment was self-reported by participants using the 11-point numerical rating scale (NRS) anchored with ‘no pain’ at 0 and ‘worst pain possible’ at 10. Disability was assessed using the 24-point Roland Morris Disability Questionnaire (RMDQ) on the day baseline and follow-up testing. An item receives a score of 1 if it is applicable to the respondent or 0 if it is not, with a total range of 0 (no disability) to 24 (severe disability) [70].

### 2.4. Sample size

Sample size for the primary study aim (i.e. to determine whether cortical reorganisation, an individual’s capacity for neuroplasticity, central sensitisation, psychosocial factors, and their possible interaction, predict LBP outcome) was initially calculated (pre study commencement) based on an assumption that the prediction model would include 17 candidate predictors, 5 ‘a priori’ interactions and 9 sociodemographic variables. Allowing for 10% loss to follow-up, a power of 80 % with a 5% level of significance and a medium effect size, a sample size of 264 participants was required. Once data collection commenced, a slower than expected rate of participant recruitment made the target sample size unachievable. On this basis, the sample size calculation for the primary aim was revised using the rule of thumb that ten subjects per variable are required to adequately power a linear regression model [14] and a minimum of five events per candidate variable is required for logistic regression analysis [57] resulting in a required sample size of 120 individuals with acute LBP. Prior to the completion of the data collection and data analysis, the UPWaRD study was registered with the Australian and New Zealand Clinical Trials Registry (ACTRN12619000002189) and the protocol for the primary study aim was published [21]. Both documents include description of the revised sample size calculation and this sample size (N=120) was achieved as planned.

### 2.5. Statistical analyses

Statistical Package for the Social Sciences software (version 27; IBM Corp) was used for all analyses in this study. Statistical significance was accepted at *P* ≤ 0.05 and all analyses were conducted on complete cases, with missing data described in **Supplemental File 1: Table S2**. First, the distribution of individual variables was inspected using histograms. Continuous data are presented as mean±standard deviation (normally distributed) or median[interquartile range] (non-normally distributed), and categorical data presented as number and percent (%).

To explore potential differences in low back pain recovery trajectories at three- and six-months, participants were divided into three sub-groups based on standardized criteria: (1) unresolved LBP if participants reported an increase or no change in pain intensity (NRS) and disability (RMDQ) from baseline, or a pain NRS score of ≥7/10 corresponding with severe pain [4]; (2) partially resolved LBP if participants reported a decrease in pain and/or disability from baseline (≥ 1-point reduction on NRS and/or RMDQ from baseline scores); or, (3) resolved LBP if participants reported no pain and disability (NRS and RMDQ = 0) at follow-up [4;25].

Comparisons were made between participants who did or did not complete follow-up, and between participants with or without LBP using independent samples t-test, non-parametric Mann-Whitney U test and Fisher’s Exact test for normally distributed, non-normally distributed and categorical data respectively. Spearman’s rank correlation coefficients were used to determine whether depression and anxiety (DASS-21), pain catastrophising (PCS) or pain self-efficacy (PSEQ) were correlated with six-month pain intensity (NRS) or disability in the UPWaRD LBP participants, with p-values adjusted for multiple comparisons using Bonferroni correction. A one-way multivariate analysis of variance (MANOVA) was used to compare differences in moderate and vigorous physical activity minutes at baseline, three-month and six-months between pain-free controls, participants with resolved LBP or participants with partially or unresolved LBP.

## 3. RESULTS

### 3.1. Participant recruitment

Between 14^th^ of April 2015 and 25^th^ July 2019, 498 participants who presented with LBP were screened and 120 participants were included in the cohort (**Figure 1**; age 39±15 years; range = 21 to 83 years, female:male sex = 59:61). Two hundred and seven participants (41.5%) were ineligible because they had chronic LBP, two participants were excluded because they had previous spinal surgery and three were excluded because physical examination by the study investigator suggested a diagnosis of lumbosacral radiculopathy. Of the 286 eligible participants, 94 (32.9%) failed to respond to contact attempts organising baseline assessment and 72 (25.2%) declined participation after reviewing the study information sheet. Baseline data were obtained on average 2.4±1.4 weeks (range 1 day to 6 weeks) after the onset of acute LBP.

**Figure 1.**
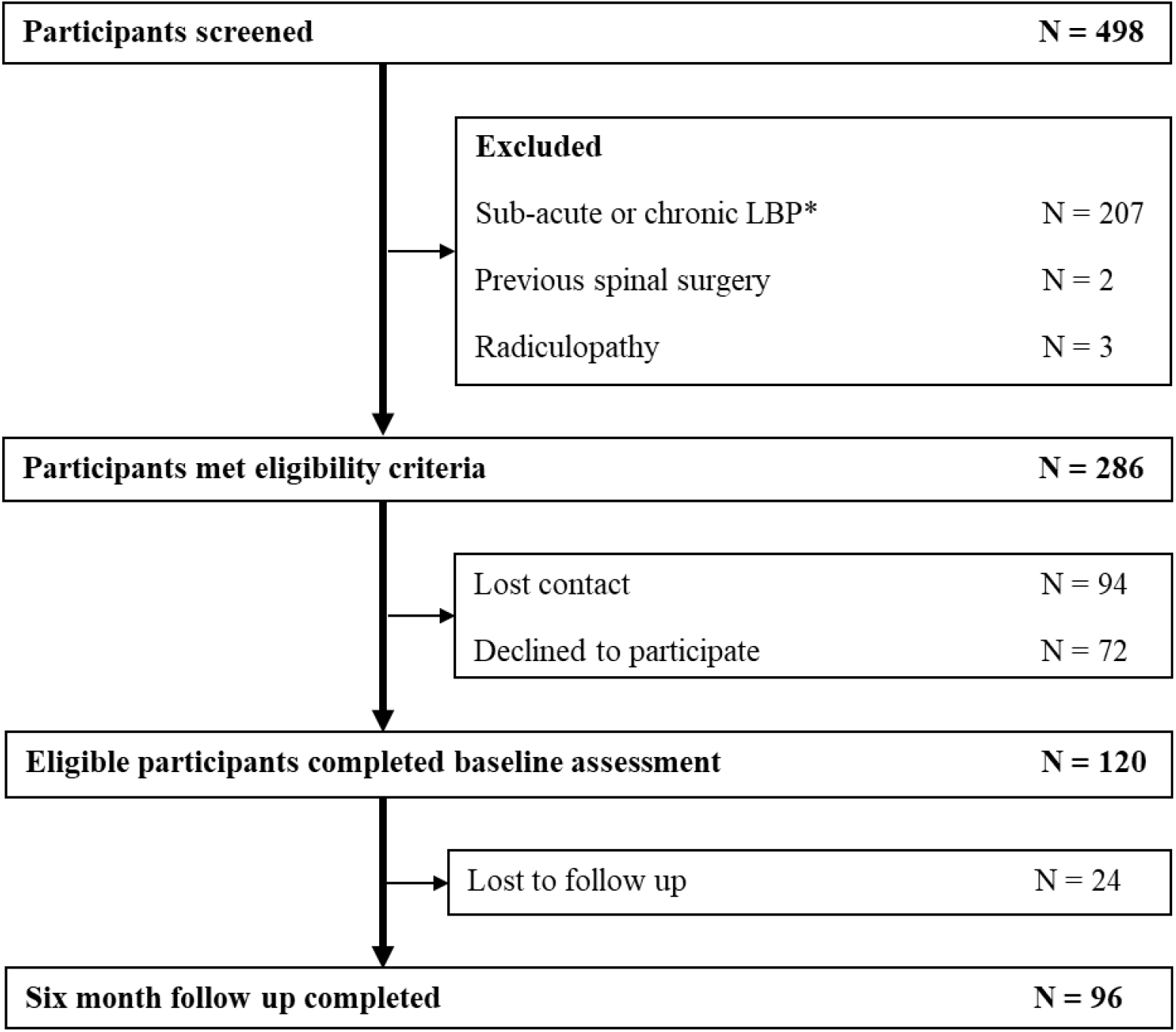
UPWaRD LBP cohort flow diagram. *defined as LBP lasting for longer than 6 weeks and/or an LBP episode preceded by a period of less than one-month without pain.

Between October 2016 and February 2019, 57 pain-free controls who reported no current or prior LBP during the 12 months preceding study entry, and with age and sex distribution similar to the UPWaRD LBP cohort were recruited (female:male sex = 28:29; age 35±14 years; range = 19 – 68, Table 2).

### 3.2. Participant attrition

Of the 120 eligible acute LBP participants who were enrolled in the study and provided baseline data, 95 participants (79%) provided outcome data at three-month follow-up and 96 participants (80%) at six-months. Missing follow-up cases were due to the participant failing to respond to multiple contact attempts to schedule their laboratory assessment within a one-month time window of their three- or six-month follow-up date. At three and six-month follow up, 15 (16%) of the 95 LBP participants and 12 (13%) of the 96 LBP participants declined assessment of all laboratory measures, respectively. These participants agreed to complete questionnaire data, and thus, remained in the cohort. **Supplemental File 1: Table S2** shows the number of participants that provided valid data for each of the three and six-month questionnaire-based items. Of the 57 control participants, follow-up was completed in 43 (75%) participants at three-months and 39 (68%) participants at six-months. Reasons for participant attrition amongst controls were as follows: i.) only consented to single laboratory testing session (N=7); ii.) withdrew from study due to intolerance of laboratory testing and/or duration of the testing protocol (N=4); iii.) no reason given (N=6); iv.) developed LBP (N=1).

Participants with higher DASS depression (*P* = <0.01), DASS anxiety (*P* = 0.03), and DASS stress (*P* = <0.01) scores and lower PSEQ (*P* = 0.02) scores were less likely to complete three-month follow-up. At six-months, participants who did not complete follow-up reported higher rates of pain affecting their work (P = 0.04), pain interference with their usual work (P = 0.03), pain interference with their walking (P = 0.04) and pain interference with their relations (P = 0.04). Higher levels of self-reported moderate physical activity time per day (P = 0.03) and lower PSEQ scores (P = 0.04) were also observed in participants who did not attend their six-month follow-up appointment (**Table 1**).

**Table 1.**
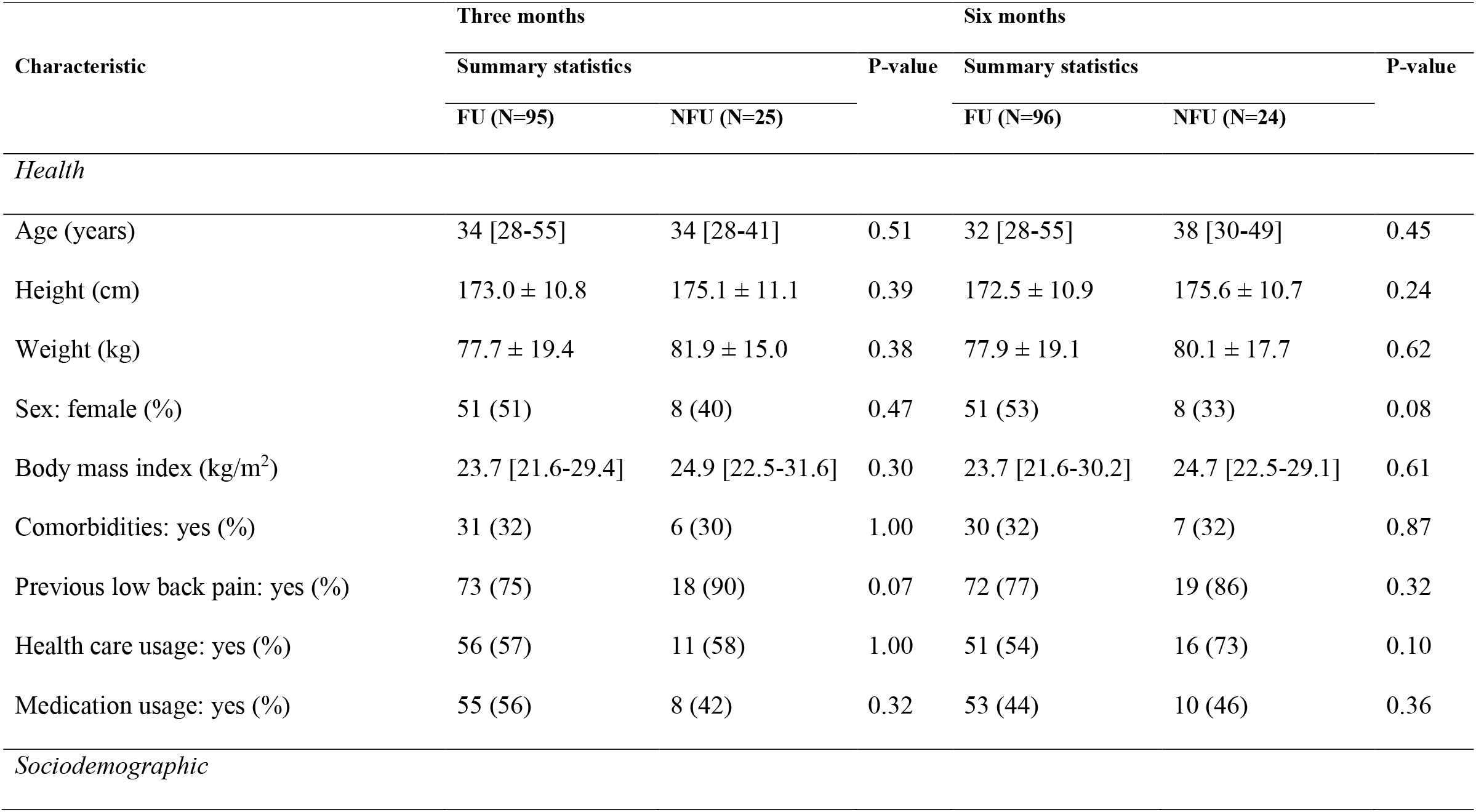
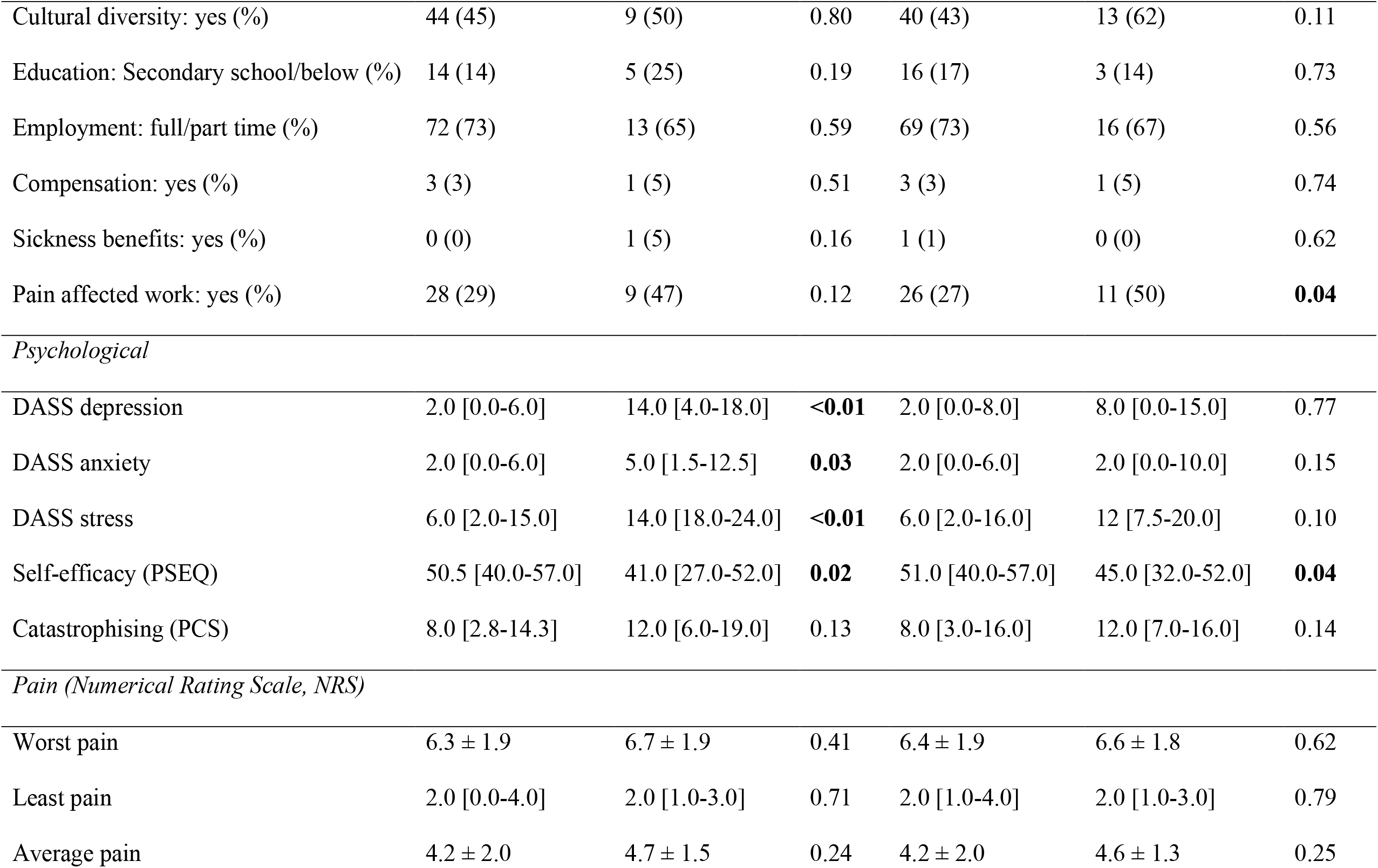
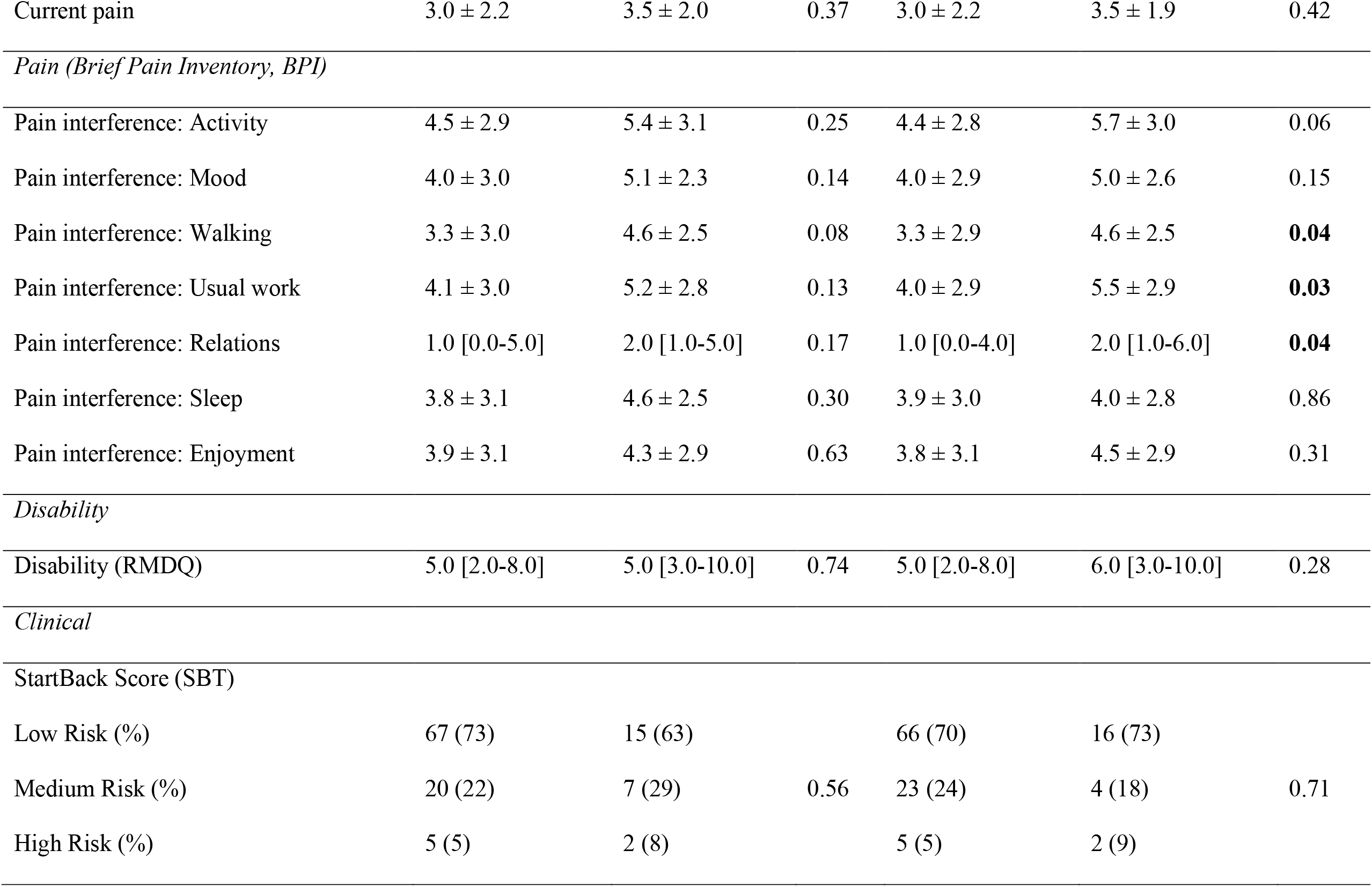
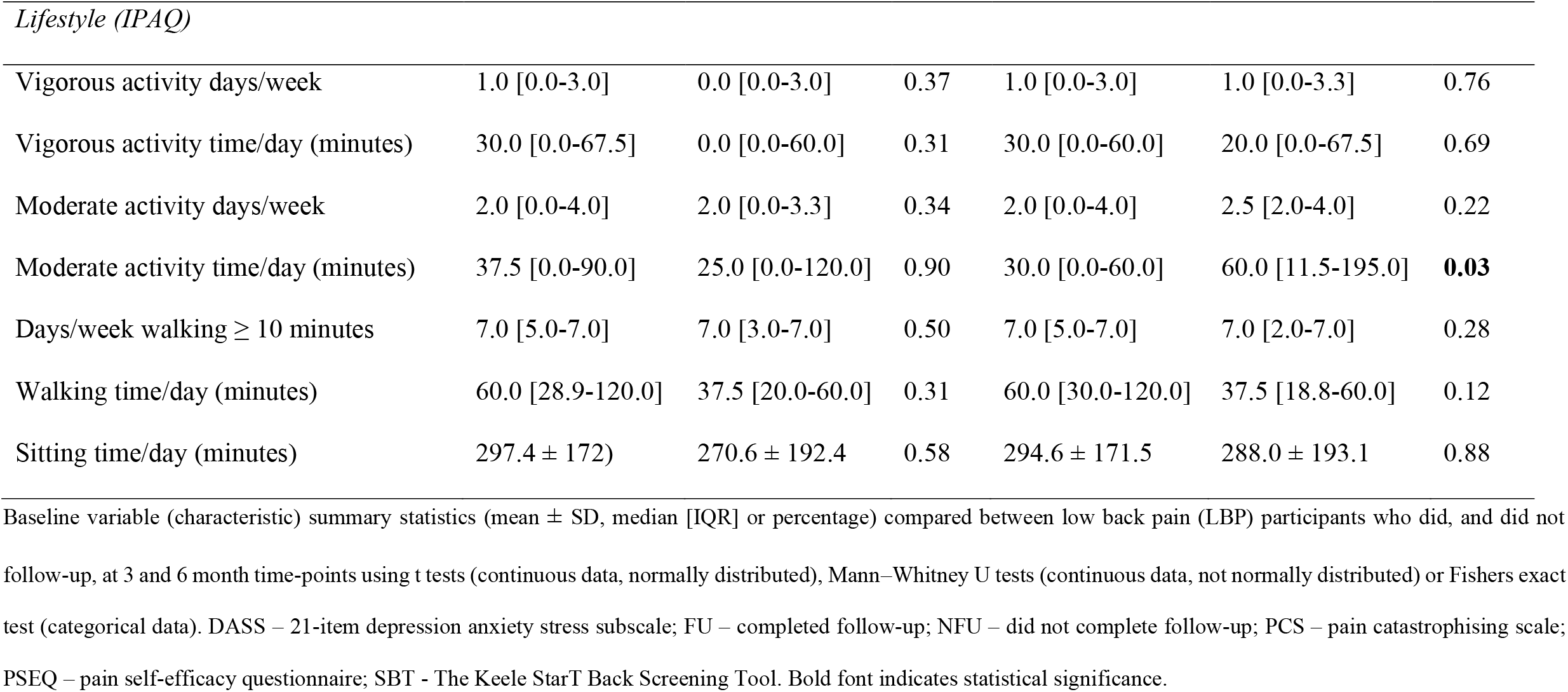
Comparison of baseline characteristics between participants with low back pain who did (FU), and did not (NFU), complete three and six-month follow-up.

| Characteristic | Three months |  | P-value | Six months |  | P-value |
| --- | --- | --- | --- | --- | --- | --- |
|  | Summary statistics |  |  | Summary statistics |  |  |
|  | FU (N=95) | NFU (N=25) |  | FU (N=96) | NFU (N=24) |  |
| *Health* |  |  |  |  |  |  |
| Age (years) | 34 [28-55] | 34 [28-41] | 0.51 | 32 [28-55] | 38 [30-49] | 0.45 |
| Height (cm) | 173.0 ± 10.8 | 175.1 ± 11.1 | 0.39 | 172.5 ± 10.9 | 175.6 ± 10.7 | 0.24 |
| Weight (kg) | 77.7 ± 19.4 | 81.9 ± 15.0 | 0.38 | 77.9 ± 19.1 | 80.1 ± 17.7 | 0.62 |
| Sex: female (%) | 51 (51) | 8 (40) | 0.47 | 51 (53) | 8 (33) | 0.08 |
| Body mass index (kg/m<sup>2</sup>) | 23.7 [21.6-29.4] | 24.9 [22.5-31.6] | 0.30 | 23.7 [21.6-30.2] | 24.7 [22.5-29.1] | 0.61 |
| Comorbidities: yes (%) | 31 (32) | 6 (30) | 1.00 | 30 (32) | 7 (32) | 0.87 |
| Previous low back pain: yes (%) | 73 (75) | 18 (90) | 0.07 | 72 (77) | 19 (86) | 0.32 |
| Health care usage: yes (%) | 56 (57) | 11 (58) | 1.00 | 51 (54) | 16 (73) | 0.10 |
| Medication usage: yes (%) | 55 (56) | 8 (42) | 0.32 | 53 (44) | 10 (46) | 0.36 |
| *Sociodemographic* |  |  |  |  |  |  |
| Cultural diversity: yes (%) | 44 (45) | 9 (50) | 0.80 | 40 (43) | 13 (62) | 0.11 |
| Education: Secondary school/below (%) | 14 (14) | 5 (25) | 0.19 | 16 (17) | 3 (14) | 0.73 |
| Employment: full/part time (%) | 72 (73) | 13 (65) | 0.59 | 69 (73) | 16 (67) | 0.56 |
| Compensation: yes (%) | 3 (3) | 1 (5) | 0.51 | 3 (3) | 1 (5) | 0.74 |
| Sickness benefits: yes (%) | 0 (0) | 1 (5) | 0.16 | 1 (1) | 0 (0) | 0.62 |
| Pain affected work: yes (%) | 28 (29) | 9 (47) | 0.12 | 26 (27) | 11 (50) | **0.04** |
| *Psychological* |  |  |  |  |  |  |
| DASS depression | 2.0 [0.0-6.0] | 14.0 [4.0-18.0] | **<0.01** | 2.0 [0.0-8.0] | 8.0 [0.0-15.0] | 0.77 |
| DASS anxiety | 2.0 [0.0-6.0] | 5.0 [1.5-12.5] | **0.03** | 2.0 [0.0-6.0] | 2.0 [0.0-10.0] | 0.15 |
| DASS stress | 6.0 [2.0-15.0] | 14.0 [18.0-24.0] | **<0.01** | 6.0 [2.0-16.0] | 12 [7.5-20.0] | 0.10 |
| Self-efficacy (PSEQ) | 50.5 [40.0-57.0] | 41.0 [27.0-52.0] | **0.02** | 51.0 [40.0-57.0] | 45.0 [32.0-52.0] | **0.04** |
| Catastrophising (PCS) | 8.0 [2.8-14.3] | 12.0 [6.0-19.0] | 0.13 | 8.0 [3.0-16.0] | 12.0 [7.0-16.0] | 0.14 |
| *Pain (Numerical Rating Scale, NRS)* |  |  |  |  |  |  |
| Worst pain | 6.3 ± 1.9 | 6.7 ± 1.9 | 0.41 | 6.4 ± 1.9 | 6.6 ± 1.8 | 0.62 |
| Least pain | 2.0 [0.0-4.0] | 2.0 [1.0-3.0] | 0.71 | 2.0 [1.0-4.0] | 2.0 [1.0-3.0] | 0.79 |
| Average pain | 4.2 ± 2.0 | 4.7 ± 1.5 | 0.24 | 4.2 ± 2.0 | 4.6 ± 1.3 | 0.25 |
| Current pain | 3.0 ± 2.2 | 3.5 ± 2.0 | 0.37 | 3.0 ± 2.2 | 3.5 ± 1.9 | 0.42 |
| *Pain (Brief Pain Inventory, BPI)* |  |  |  |  |  |  |
| Pain interference: Activity | 4.5 ± 2.9 | 5.4 ± 3.1 | 0.25 | 4.4 ± 2.8 | 5.7 ± 3.0 | 0.06 |
| Pain interference: Mood | 4.0 ± 3.0 | 5.1 ± 2.3 | 0.14 | 4.0 ± 2.9 | 5.0 ± 2.6 | 0.15 |
| Pain interference: Walking | 3.3 ± 3.0 | 4.6 ± 2.5 | 0.08 | 3.3 ± 2.9 | 4.6 ± 2.5 | **0.04** |
| Pain interference: Usual work | 4.1 ± 3.0 | 5.2 ± 2.8 | 0.13 | 4.0 ± 2.9 | 5.5 ± 2.9 | **0.03** |
| Pain interference: Relations | 1.0 [0.0-5.0] | 2.0 [1.0-5.0] | 0.17 | 1.0 [0.0-4.0] | 2.0 [1.0-6.0] | **0.04** |
| Pain interference: Sleep | 3.8 ± 3.1 | 4.6 ± 2.5 | 0.30 | 3.9 ± 3.0 | 4.0 ± 2.8 | 0.86 |
| Pain interference: Enjoyment | 3.9 ± 3.1 | 4.3 ± 2.9 | 0.63 | 3.8 ± 3.1 | 4.5 ± 2.9 | 0.31 |
| *Disability* |  |  |  |  |  |  |
| Disability (RMDQ) | 5.0 [2.0-8.0] | 5.0 [3.0-10.0] | 0.74 | 5.0 [2.0-8.0] | 6.0 [3.0-10.0] | 0.28 |
| Low Risk (%) | 67 (73) | 15 (63) |  | 66 (70) | 16 (73) |  |
| Medium Risk (%) | 20 (22) | 7 (29) | 0.56 | 23 (24) | 4 (18) | 0.71 |
| High Risk (%) | 5 (5) | 2 (8) |  | 5 (5) | 2 (9) |  |

| *Lifestyle (IPAQ)* |  |  |  |  |  |  |
| --- | --- | --- | --- | --- | --- | --- |
| Vigorous activity days/week | 1.0 [0.0-3.0] | 0.0 [0.0-3.0] | 0.37 | 1.0 [0.0-3.0] | 1.0 [0.0-3.3] | 0.76 |
| Vigorous activity time/day (minutes) | 30.0 [0.0-67.5] | 0.0 [0.0-60.0] | 0.31 | 30.0 [0.0-60.0] | 20.0 [0.0-67.5] | 0.69 |
| Moderate activity days/week | 2.0 [0.0-4.0] | 2.0 [0.0-3.3] | 0.34 | 2.0 [0.0-4.0] | 2.5 [2.0-4.0] | 0.22 |
| Moderate activity time/day (minutes) | 37.5 [0.0-90.0] | 25.0 [0.0-120.0] | 0.90 | 30.0 [0.0-60.0] | 60.0 [11.5-195.0] | **0.03** |
| Days/week walking ≥ 10 minutes | 7.0 [5.0-7.0] | 7.0 [3.0-7.0] | 0.50 | 7.0 [5.0-7.0] | 7.0 [2.0-7.0] | 0.28 |
| Walking time/day (minutes) | 60.0 [28.9-120.0] | 37.5 [20.0-60.0] | 0.31 | 60.0 [30.0-120.0] | 37.5 [18.8-60.0] | 0.12 |
| Sitting time/day (minutes) | 297.4 ± 172) | 270.6 ± 192.4 | 0.58 | 294.6 ± 171.5 | 288.0 ± 193.1 | 0.88 |

Baseline variable (characteristic) summary statistics (mean ± SD, median [IQR] or percentage) compared between low back pain (LBP) participants who did, and did not follow-up, at 3 and 6 month time-points using t tests (continuous data, normally distributed), Mann–Whitney U tests (continuous data, not normally distributed) or Fishers exact test (categorical data). DASS – 21-item depression anxiety stress subscale; FU – completed follow-up; NFU – did not complete follow-up; PCS – pain catastrophising scale; PSEQ – pain self-efficacy questionnaire; SBT - The Keele StarT Back Screening Tool. Bold font indicates statistical significance.

**Table 2.** UPWaRD LBP cohort pain and disability outcomes at three and six-month follow up.

| **Classification** | **Three months: N (%)** | **Six months: N (%)** | **P-value** |
| --- | --- | --- | --- |
| Unresolved recurrent or chronic LBP | 16 (16.8) | 12 (12.5) | 0.21 |
| Partially resolved recurrent or chronic LBP | 57 (60.0) | 60 (62.5) | **0.01** |
| Resolved | 22 (23.2) | 24 (25.0) | **<0.001** |

Summary statistics are number (%), compared between three- and six-month time points using Fishers Exact test.
LBP outcome within the UPWaRD Cohort was dichotomised at three and six-months using standardized criteria defined in Section 2.5.

### 3.3. Pain and disability recovery trajectories

Overall, NRS scores of pain intensity for participants with LBP decreased (P=<0.001) from 4.3±1.9 at baseline to 2.3±2.3 at three-months, remaining stable at six-months (2.3±2.2). Disability scores (RMDQ) decreased (P=<0.001) from a median score of 5.0 (IQR = 2.0 – 8.3) at baseline to a median score of 2.0 (IQR = 0.0 – 5.0) at three-months, and a median score of 1.0 (IQR = 0.0 – 4.0) at six-months. Reporting unresolved LBP at three-months was not significantly associated with experiencing unresolved LBP at six-months (P = 0.21). Conversely, partially resolved (P = 0.01) and resolved (P = <0.001) LBP status at three-months was significantly associated with six-month partially resolved and resolved LBP status, respectively. Twenty-four (25.0%) LBP participants were completely recovered and sixty (62.5%) were partially recovered after six months. Twelve (12.5%) participants LBP was unresolved at six-months (**Table 2**).

### 3.4. Health-related characteristics

Compared to controls, LBP participants were slightly older, had a higher body mass index (BMI), a higher prevalence of comorbidities and higher medication usage (**Table 3**). The most reported comorbidities amongst participants with LBP were depression/anxiety (N=12, 29.3%), hypertension (N=9, 22.0%) and asthma (N=5, 12.2%). Amongst controls, six comorbidities were self-reported: vision impairment (N=1), hypothyroidism (N=1), osteoporosis (N=1), prolactinoma (N=1), mild depression/anxiety not requiring intervention (N=1), and heart disease (N=1). The most frequently used medication within the control group was a contraceptive (N=4). Types of health care utilised by LBP participants were allied health (N=59, 50.4%), general practitioners (N=30, 25.6%), diagnostic tests (N=13, 11.1%), and specialist physicians (N=5, 4.3%). During the follow-up period, three (2.6%) participants presented to their local emergency department because of their LBP but none were admitted to hospital. Amongst participants experiencing an acute episode of LBP, fifty-five (46.6%) did not use any medication and two (1.7%) did not specify their medication use. Eighteen (15.3%) used nonsteroidal anti-inflammatories and 19 (16.1%) used acetaminophen. Seven LBP participants (5.9%) were prescribed opioids and three (2.5%) were prescribed benzodiazepines. Nine LBP participants (7.6%) were taking anti-depressant medication for the management of co-existing depressive symptoms. Three LBP participants were prescribed an anti-convulsant (2.5%). No LBP participants in the UPWaRD cohort received an epidural steroid injection. Thirty-three participants with LBP were taking medication not related to pain (e.g. anti-hypertensive or oral contraceptives).

**Table 3.** Baseline demographic and health-related characteristics of the UPWaRD cohort

| Health-related characteristic | Summary statistics |  |  |
| --- | --- | --- | --- |
|  | Low back pain (N=120) | Control (N=57) | P-value |
| Age (years) | 34 [28-53] | 31 [25-40] | **0.02** |
| Height (cm) <sup>\$</sup> | 173.1 ± 10.9 | 170.6 ± 8.2 | 0.10 |
| Weight (kg) <sup>\$</sup> | 78.3 ± 18.8 | 69.0 ± 13.7 | **<0.001** |
| Sex (female, %) | 59 (49) | 28 (49) | 0.56 |
| Comorbidities (yes, %) | 37 (32) | 6 (11) | **<0.01** |
| Previous LBP (yes, %) | 91 (78) | 2 (4.0) | **<0.001** |
| Health care usage (yes, %) | 67 (57) | NA | NA |
| Medication usage (yes, %) | 63 (53) | 12 (21) | **<0.001** |
| Body Mass Index (kg/m<sup>2</sup>) | 24.2 [21.7-29.8] | 22.5 [21.2-25.8] | **0.01** |

Summary statistics (mean ± SD, median [IQR] or percentage) compared between low back pain and control participants using *t* tests (continuous data, normally distributed), Mann-Whitney *U* tests (continuous data, not normally distributed) or Fishers Exact test (categorical data). <sup>\$</sup> Welch's *t*-test was performed. Bold font indicates statistical significance

### 3.5. Sociodemographic characteristics

Fifty-three (46.1%) LBP participants and 30 (56.6%) pain-free controls identified as culturally diverse. Only one participant with LBP was receiving sickness benefits (0.9%) at the time of baseline testing and four (3.4%) were receiving compensation related to their LBP. Thirty-seven (31.6%) LBP participants reported pain that was affecting their occupation. **Table 4** outlines the education and occupational status of the UPWaRD cohort.

**Table 4.**
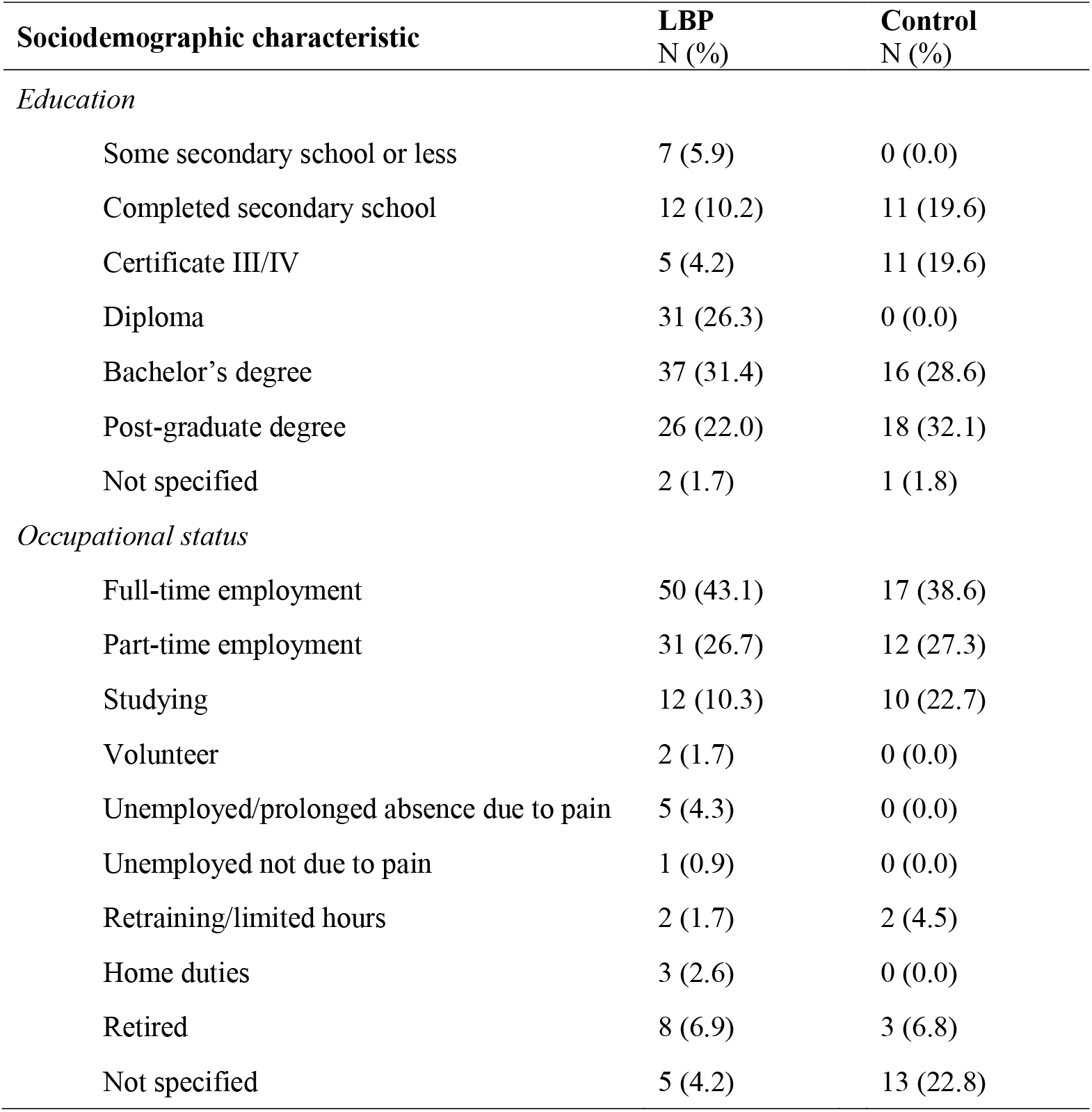
Education and occupational status of participants enrolled in the UPWaRD study

| **Sociodemographic characteristic** | **LBP**<br>**N (%)** | **Control**<br>**N (%)** |
| --- | --- | --- |
| *Education* |  |  |
| Some secondary school or less | 7 (5.9) | 0 (0.0) |
| Completed secondary school | 12 (10.2) | 11 (19.6) |
| Certificate III/IV | 5 (4.2) | 11 (19.6) |
| Diploma | 31 (26.3) | 0 (0.0) |
| Bachelor's degree | 37 (31.4) | 16 (28.6) |
| Post-graduate degree | 26 (22.0) | 18 (32.1) |
| Not specified | 2 (1.7) | 1 (1.8) |
| *Occupational status* |  |  |
| Full-time employment | 50 (43.1) | 17 (38.6) |
| Part-time employment | 31 (26.7) | 12 (27.3) |
| Studying | 12 (10.3) | 10 (22.7) |
| Volunteer | 2 (1.7) | 0 (0.0) |
| Unemployed/prolonged absence due to pain | 5 (4.3) | 0 (0.0) |
| Unemployed not due to pain | 1 (0.9) | 0 (0.0) |
| Retraining/limited hours | 2 (1.7) | 2 (4.5) |
| Home duties | 3 (2.6) | 0 (0.0) |
| Retired | 8 (6.9) | 3 (6.8) |
| Not specified | 5 (4.2) | 13 (22.8) |

### 3.6. Psychological characteristics

DASS depression scores were higher at baseline in acute LBP participants compared with pain-free controls (*P* = 0.01). Although the median total DASS-21 scores appeared higher at baseline in the acute LBP participants compared with pain-free controls, the distributions overlapped and did not differ significantly (*P* = 0.13; **Table 5**). PCS and PSEQ scores were not obtained at baseline from pain-free participants however the median scores for these measurements amongst LBP participants are presented in **Table 5**.

**Table 5.** Baseline psychological characteristics of the UPWaRD cohort

| **Psychological characteristic** | **Summary statistics** |  |  |
| --- | --- | --- | --- |
|  | **Low back pain (N=120)** | **Control (N=57)** | **P-value** |
| DASS total | 16.0 [4.0-28.0] | 10.0 [4.0-22.0] | 0.13 |
| DASS depression item | 2.0 [0.0-10.0] | 2.0 [0.0-4.0] | **0.01** |
| DASS anxiety item | 2.0 [0.0-6.0] | 2.0 [0.0-4.0] | 0.12 |
| DASS stress item | 8.0 [2.0-16.0] | 8.0 [4.0 – 12.0] | 0.37 |
| PCS | 8.0 [3.0-15.5] | NA | NA |
| PSEQ | 48.0 [37.5 – 56.0] | NA | **NA** |

Summary statistics (median [IQR]) compared between low back pain and control participants using Mann-Whitney *U* tests (continuous data, not normally distributed). Bold font indicates statistical significance. DASS – 21-item depression anxiety stress subscale; PCS – pain catastrophising scale; PSEQ – pain self-efficacy questionnaire; NRS – numerical rating scale; RMDQ – Roland Morris disability questionnaire.

**Table 6 reports** correlations between psychological variables of interest and six-month pain (NRS) and disability (RMDQ) in the LBP cohort (NRS). All psychological variables at baseline were significantly correlated (*P* = <0.05) with six-month pain intensity and disability.

**Table 6.**
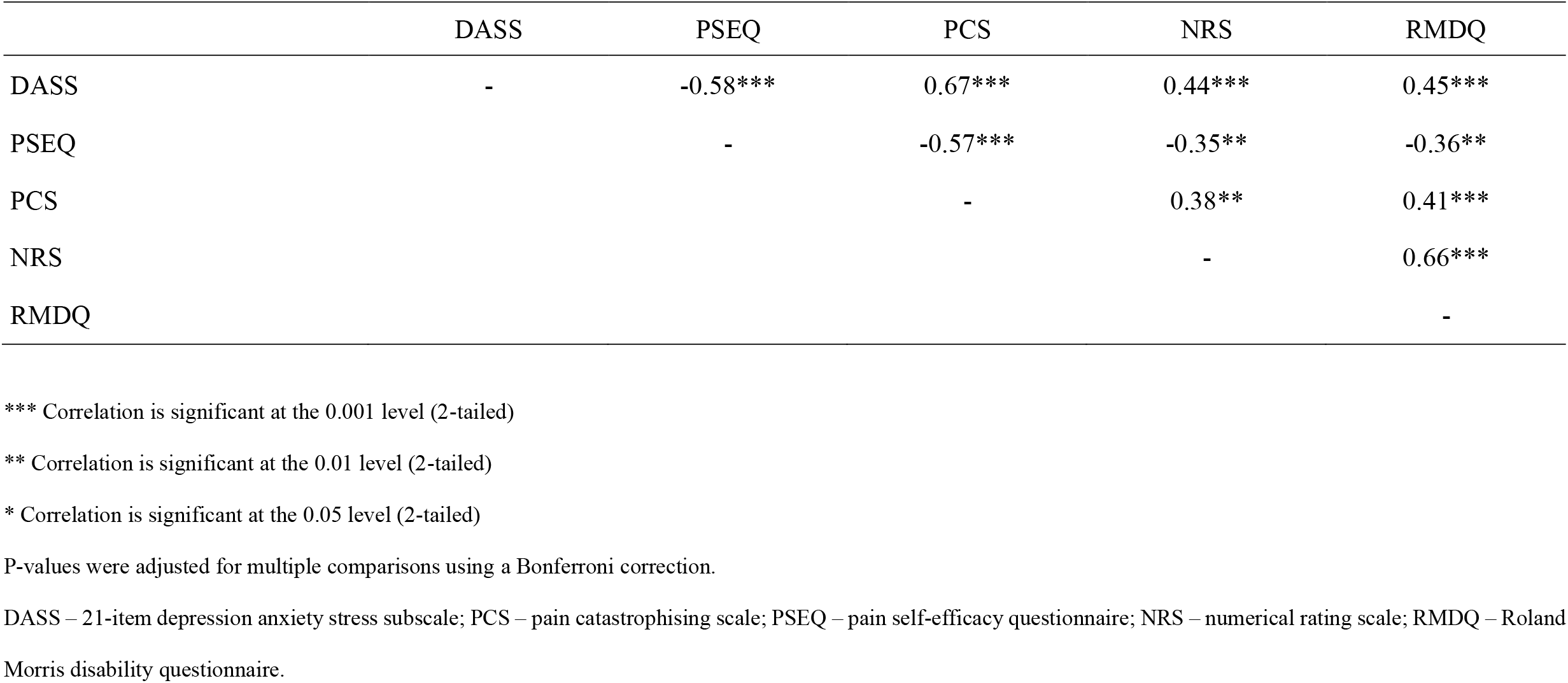
Spearman’s correlation coefficients between measures of baseline psychological status and six-month pain and disability

### 3.7. Lifestyle characteristics

Compared to pain-free controls, participants in the UPWaRD LBP cohort engaged in lower levels of vigorous and moderate physical activity the week preceding their first laboratory session (*P =* <0.05; **Table 7**). Amongst the complete cases, there was no difference in moderate physical activity minutes between groups (controls, resolved LBP, partially or unresolved LBP) at three-month follow-up (F_6, 176_ = 0.96, *P* = 0.45; Wilks’ Lambda = 0.94, Partial Eta^2^ = 0.03), and a similar result was observed at six-month follow-up (F_6, 174_ = 1.25, *P* = 0.28; Wilks’ Lambda = 0.92, Partial Eta^2^ = 0.04). Vigorous physical activity minutes amongst complete cases also did not differ between groups at three-month (F_6, 192_ = 0.85, *P* = 0.53; Wilks’ Lambda = 0.95, Partial Eta^2^ = 0.03), or at six-months (F_6, 192_ = 0.86, *P* = 0.52; Wilks’ Lambda = 0.95, Partial Eta^2^ = 0.03).

**Table 7.** Baseline physical activity levels of the UPWaRD cohort based on the International Physical Activity Questionnaire (IPAQ)

| Lifestyle-related characteristic | Summary statistics |  |  |
| --- | --- | --- | --- |
|  | Low back pain (N=120) | Control (N=57) | P-value |
| Vigorous. activity days/week | 1.0 [0.0-3.0] | 2.0 [1.0-4.0] | **0.01** |
| Vigorous. activity time/day (minutes) | 30.0 [0.0-60.0] | 60.0 [20.0-90.0] | **0.01** |
| Moderate activity days/week | 2.0 [0.0-4.0] | 3.0 [2.0-5.0] | **0.01** |
| Moderate activity time/day (minutes) | 30.0 [0.0-90.0] | 60.0 [30.0-120.0] | **0.02** |
| Days/week walking ≥ 10 minutes | 7.0 [4.0-7.0] | 7.0 [5.0-7.0] | 0.15 |
| Walking time/day (minutes) | 45.0 [25.0-120.0] | 60.0 [30.0-120.0] | 0.16 |
| Sitting time/day (minutes) | 293.4 ± 174.8 | 291.0 ± 205.1 | 0.94 |

Summary statistics are mean ± SD or median [IQR], compared between low back pain and control participants using participants using *t* tests (continuous data, normally distributed) or Mann-Whitney *U* tests (continuous data, not normally distributed). Bold font indicates statistical significance.

## 4. DISCUSSION

This cohort profile describes the sample characteristics of 120 adults experiencing an episode of acute LBP and 57 pain-free control participants recruited for the UPWaRD study. This cohort provides a unique opportunity to investigate changes occurring across several neurobiological systems, and their interactions with heritable and environmental traits, during the transition from acute to chronic or recurrent LBP.

### 4.1. Cohort baseline characteristics

LBP is a heterogenous condition [20] and contributors to pain chronicity and disability are multifactorial [15]. LBP participants in the UPWaRD cohort, were slightly older, had a higher average BMI and participated in lower levels of vigorous and moderate physical activity the week preceding baseline testing than the pain-free controls recruited for this study. Although this might be expected for individuals with pain, a recent systematic review including individuals free from chronic-LBP at study inception, suggests lower levels of moderate (1–3 times per week), or vigorous/high (≥3–4 times per week) leisure physical activity may increase the risk of developing chronic LBP [46]. A significant causal relationship has recently been identified between BMI and back pain development [11].

An important finding of the cohort profile presented here was that over 50% of the UPWaRD LBP cohort utilized at least one form of health care because of their LBP episode, most commonly, allied health (e.g. physiotherapist, chiropractor) or general practitioners. Notably, 11% of the UPWaRD LBP cohort underwent diagnostic imaging for their acute LBP episode, 6% received opioids for management of their LBP symptoms and 4% received a specialty consultation (e.g. spinal surgeon). Routine use of diagnostic imaging, opioid medication and specialist consultation in the absence of serious pathology is not recommended for acute LBP [39]. As all participants in the UPWaRD cohort were carefully screened for the presence of serious pathology and signs of lumbosacral radiculopathy, this finding is likely to represent care that is discordant with current clinical practice guidelines. The observation of discordant care is consistent with studies of individuals with acute LBP presenting to Australian emergency departments [31]. A recent prospective cohort study identified a linear relationship between guideline discordant care and increased risk of transition to chronicity [50]. International guidelines consistently recommend general practitioners provide advice, education, reassurance, and simple analgesics, when necessary to manage acute, non-specific LBP [54]. Recently, non-pharmacologic interventions, such as heat, massage, acupuncture, or spinal manipulation have also been recommended as first-line treatment options [39].

Previous research has linked psychological risk factors with the transition from acute to chronic LBP [30;43]. Psychological risk factors (i.e. depression, anxiety and stress, pain catastrophising and pain self-efficacy beliefs) assessed in the UPWaRD acute LBP cohort at baseline were comparable to that of the pain-free participants, a finding that has been observed in previous comparable cohorts [42]. However, amongst the UPWaRD acute LBP participants, higher levels of depression, anxiety and stress, higher pain catastrophising and lower pain self-efficacy at baseline were correlated with higher six-month pain intensity and disability (**Table 6**). Systematic reviews of thirteen LBP cohorts report similar findings, with depression and catastrophizing consistently identified as significant risk factors for poor LBP outcome [44;60].

### 4.2. Pain and disability recovery trajectories

On average, LBP participants included in the UPWaRD cohort demonstrated a significant reduction in pain and disability between baseline and three-months, yet no significant change in pain intensity and disability from three-month to six-month assessment. This is typical of LBP studies. A meta-analysis of 33 discrete cohorts identified a comparable recovery trajectory [5]. Further, the UPWaRD LBP cohort reported similar recovery rates to previous acute LBP cohorts [27]. At six months, 12 (12.5%) LBP participants in the UPWaRD cohort reported worse pain and disability from baseline or severe pain (NRS≥7), 60 (62.5%) participants reported less pain and disability compared to baseline, and 24 (25%) participants reported no pain or disability. In the cohort described by Klyne and colleagues [27], 15 (15.5%) participants reported worse or severe LBP, 66 (68.0%) participants reported less pain and disability and 16 (16.5%) participants reported no pain or disability at six-month follow-up. Similar rates of ongoing LBP at six-month follow-up have been reported in other LBP cohorts [3;34;35]. However, individuals in the UPWaRD LBP cohort reported lower levels of disability at 6-months follow-up when compared with other chronic LBP cohorts [37;38].

### 4.3. Methodological issues

The baseline characteristics presented here provide a foundation for future longitudinal analyses, however, the UPWaRD study is not without limitations. Although missing data are inevitable in longitudinal trials, the presence of incomplete cases does represent a threat to the depth of the results. The UPWaRD cohort reports similar rates of missing data to most recent prospective cohort studies examining biological risk factors during an acute LBP episode [27;35;56]. Most missing data in this cohort occurred after the first laboratory session, and many baseline characteristics, with some exceptions, were similar between those who did and did not return for follow-up. Study attrition was likely due to inclusion of a high burden of laboratory measures that some participants found difficult to tolerate, and the time-commitment involved in the study. In this cohort, individuals who were lost to follow up at three- or six-months reported, at baseline, higher levels of depression, anxiety, stress, and pain catastrophising, higher pain interference, higher levels of moderate physical activity, and occupational difficulties due to pain. Future longitudinal cohort studies might benefit from considering this finding and implementing targeted, innovative methods to reduce attrition in participants with similar baseline characteristics.

Difficulties were experienced with recruitment, highlighted by the revised sample size and time taken to recruit the required number of LBP participants. Similar difficulties with recruitment have been reported by other groups conducting experimental LBP cohort studies [27;35]. Cohort studies conducted alongside randomized trials of new treatments appear to have greater recruitment success [50] and this may be an important consideration for future LBP cohort study designs.

Another important limitation to consider is that pain and disability outcome measures for the UPWaRD LBP cohort were assessed over the week preceding three and six-month follow-up assessment. Consequently, it is not possible to determine whether the presence of pain and disability at six-months follow-up reflects chronic LBP (i.e. pain that had persisted since the acute episode) or chronic recurrent LBP (i.e. a new episode of LBP following a pain-free period). This is acknowledged in our classification of the presence of LBP at three- and six-month follow up (i.e. chronic or recurrent LBP). More frequent assessment of pain and disability over the course of the follow-up period (e.g. weekly/second weekly would allow evaluation of differing recovery trajectories [6;25].

### 4.4. Conclusion

This cohort profile reports baseline characteristics of the UPWaRD LBP and pain-free control cohort. Overall, the UPWaRD LBP cohort represents a generalisable sample of participants experiencing an acute episode of LBP within the community, many of whom seek and utilise treatment. Psychological risk factors (i.e. higher depression, anxiety and stress, higher pain catastrophising and lower pain self-efficacy) assessed during acute LBP were correlated with higher pain and disability at six-months. Participants experiencing acute LBP were older, had a higher BMI and participated in lower levels of moderate and vigorous physical activity during an acute LBP episode compared with pain-free control participants. Participants who did not complete follow-up at three- and six-months had higher psychological distress, higher pain interference, higher levels of moderate physical activity, and reported occupational difficulties due to pain.

## Data Availability

All data produced in the present study are available upon reasonable request to the authors.

