## Supplemental File 1 for "The Understanding persistent Pain Where it ResiDes study of low back pain cohort profile"

**Table S1. Detailed description of measures collected in the UPWaRD study.**

| Measure | Description | Assessed in | | Units/range |
|---|---|---|---|---|
| | | LBP | Control | |
| *Health* | | | | |
| Age, height $^{\Delta}$, weight $^{\Delta}$, sex* | Self-reported age, height, weight, sex | ✓ | ✓ | Years, cm, kg, male/female |
| BMI $^{\Delta}$ | Weight (kg) divided by the squared height (cm) | ✓ | ✓ | Numerical |
| Comorbidities* $^{\Delta}$ | Self-selected comorbid conditions other than LBP from a list (including "other") | ✓ | ✓ | Yes/no, type |
| Health care usage $^{\Delta}$ | Self-reported health care usage from a list including general practitioner, medical specialist, health professionals other than doctors, emergency department, hospital admission and diagnostic tests | ✓ | ✓ | Frequency, type |
| Medication usage $^{\Delta}$ | Self-reported medication usage | ✓ | ✓ | Dosage, frequency, type |
| Previous LBP* | Self-reported previous incidence(s) of LBP | ✓ | ✓ | Yes/no |
| *Sociodemographic* | | | | |

| | | | | |
|---|---|---|---|---|
| Cultural diversity* Δ | Each participant was asked the question: “How do you define your identity, in ethnic or cultural terms?” If the participant identified a cultural or ethnic background other than “English”, “Caucasian” or “Australian” they were considered culturally and linguistically diverse for the purpose of this study. | ✓ | ✓ | Type |
| Education level* Δ | Self-selected highest education level from a list (e.g., primary school, completed secondary school, post-graduate degree) | ✓ | ✓ | Type |
| Employment status Δ | Self-selected employment status from a list (e.g., full-time paid employment, studying, retired) | ✓ | ✓ | Type |
| Impending compensation Δ | Self-reported impending or current compensation case related to the LBP episode | ✓ | | Yes/no, type |
| Sickness benefits Δ | Self-reported sickness benefits associated with the participants’ LBP episode | ✓ | | Yes/no |

| | | | | |
|---|---|---|---|---|
| Pain affected work [Δ] | Self-reported pain affected work hours or whether pain affects the type of work the respondent can complete | ✓ | | Yes/no |
| *Psychological* | | | | |
| 21-item Depression, anxiety and stress subscale (DASS-21 [15]) | Questionnaire: evaluates symptoms of depression, anxiety and tension-stress. Consists of 21 items with responses quantified on a four-point Likert scale ranging from 0 ("not at all") to 3 ("applied to me very much, or most of the time"). Yields a total score as well as three subscale scores: DASS-depression (low positive affect), DASS-anxiety (psychological hyper-arousal) and DASS-stress (e.g., tension or irritability) | ✓ | ✓ | 0-63: ↑ score = ↑ distress along the 3 axes of depression, anxiety and stress<br>Subscales: depression (0-21; ≥ 11 = severe), anxiety (0-21; ≥ 8 = severe), stress (0-21; ≥ 13 = severe). |
| Pain self-efficacy questionnaire (PSEQ [16]) | Questionnaire: evaluates an individual's confidence in their ability to perform a range of functional activities while in pain. Consists of 10 items. Respondents rate how confident | ✓ | | 0-60: ↑ score = ↑ self-efficacy beliefs |

| | | | |
|---|---|---|---|
| | they are in performing each item using a seven-point Likert scale | | |
| Pain catastrophising scale (PCS [22]) | Questionnaire: evaluates thoughts and feelings related to catastrophic cognitions when in pain. Consists of 13 items with responses quantified on a five-point Likert scale ranging from 0 (“not at all”) to 4 (“all the time”). Yields a total score as well as three subscales: magnification (3 items), rumination (4 items) and helplessness (6 items) | ✓ | 0–52: ↑score = ↑pain catastrophizing Subscales: magnification (0–12), rumination (0–16), helplessness (0–24) |
| *Clinical* | | | |
| The Keele StarT Back Screening Tool (SBT [10])* Δ | Questionnaire: The SBT is a brief, validated tool, designed to screen patients presenting to primary care with acute LBP. Respondents select if they agree or disagree with the first 8-items (e.g., “my back pain spread down my leg(s) at some point in the last 2 weeks”), then rate the overall bothersomeness of their LBP using a five-point Likert scale from (“0 = not at all”) to 4 (“extremely”) | ✓ | Respondents reporting a score of 0-3 are classified as low-risk and those reporting scores of ≥ 4 overall as medium-risk. Respondents are considered at high-risk |

| | | | | |
|---|---|---|---|---|
| | | | | of a worse outcome if they score 4 or 5 in the distress subscale score (questions 5 – 9) |
| *Neurophysiological* | | | | |
| Sensory evoked potentials (electroencephalography recording: SEPs) | Laboratory measure: SEPs were assessed based on our previous work demonstrating reliability of this measure in healthy participants [6]. Participants were seated comfortably in a chair with eyes closed. Electroencephalographic SEPS were recorded using gold plated cup electrodes positioned over the primary sensory cortex contralateral to the side of worst pain for LBP participants, or contralateral to the dominant hand in healthy controls, and referenced to Fz using the International 10/20 System [11]. A constant current stimulator delivered two blocks of 500 non-noxious electrical stimuli through a single bipolar electrode | ✓ | ✓ | Latency (mS), Area under the rectified curve (µV) |

| | | | | |
|---|---|---|---|---|
| | positioned 3cm lateral to the L3 spinous process, ipsilateral to the side of the worst LBP, or dominant hand for healthy controls. Individual SEP traces were manually inspected and averaged for analysis. Distinct SEP components are thought to reflect sensory afferent processing within the human cortex and the area under the rectified curve for each component was a candidate predictor reported ‘a priori’ in the study protocol ($N_{80}$ – primary sensory cortex excitability, $N_{150}$ – secondary sensory cortex excitability, $P_{260}$ – anterior cingulate cortex excitability [2;8;9;12]) | | | |
| Corticomotor excitability | Laboratory measure: The corticomotor response to transcranial magnetic stimulation (TMS) was assessed using an established mapping paradigm and based on our previous work [4;12;17;19;20;23]. Participants sat comfortably in a chair and electrodes were placed on the paraspinal muscles 3 cm lateral to the spinous process of L3 and 1 cm lateral to | ✓ | ✓ | Map volume ($cm^2$)<br>Centre of gravity (cm) |

the spinous process of L5 (Noraxon USA Inc, Arizona, USA). Participants were fitted with a tight-fitting cap, marked with a 6 x 7 cm grid oriented to the vertex. Single-pulse, monophasic stimuli (Magstim 200 stimulator/7 cm figure-of-eight coil; Magstim Co. Ltd. Dyfed, UK) was then delivered over M1 contralateral to the side of the worst LBP, starting at the vertex. For healthy controls, M1 contralateral to the dominant hand was stimulated. Five stimuli were delivered over each site on the grid with an inter-stimulus interval of 6 s at 100% of maximum stimulator output. Participants maintained activation of their paraspinal extensor muscles to 20 ± 5% of their EMG recorded during a maximum voluntary contraction throughout the stimulation with constant feedback of real-time EMG displayed on a monitor. Parameters calculated from the normalised motor cortical maps are described in the study protocol [12].

| | | | | |
|---|---|---|---|---|
| *Biological* | | | | |
| Serum cytokines [Δ] | Laboratory measure: Serum concentrations of IL-1β, IL-2, IL-4, IL-6, IL-8. IL-10, IL-15, TNF, CRP, TGF-β1. Peripheral venous blood was drawn, clotted (30 min, room temperature), and separated by centrifugation (2500 rpm, 15 min). Serum samples were pipetted into 50 µL aliquots and stored at -80°C until analysis. After thawing, concentrations of each biomarker were determined using “high-sensitive” enzyme-linked immunosorbent assays (ELISA, Protein Simple, CA, USA). Samples were loaded into the cartridge according to a standard procedure provided by the manufacturers and immunoassay scans processed with no user activity. Built in cartridge limits of detection for each biomarker were as follows: (1) IL-1β: 0.064 pg/ml; (2) IL-2: 0.18 pg/ml; (3) IL-4: 0.16 pg/ml; (4) IL-6: 0.26 pg/ml; (5) IL-8: 0.08 pg/ml; (6) IL-10: 0.14 pg/ml; (7) 1L-15: 0.19 | ✓ | ✓ | pg/mL |

| | | | | |
|---|---|---|---|---|
| | pg/ml; (8) TNF: 0.278 pg/ml; (9) CRP: 1.24 pg/ml; (10) TGF-β1: 5.29 pg/ml. Zero was allocated for values below the reported sensitivity of the test. | | | |
| Brain derived neurotrophic factor (BDNF) genotype and serum concentration* | Laboratory measure: Buccal swabs were taken on the day of baseline testing (Isohelix DNA Isolation Kit) and immediately frozen and stored at −80°C. Genomic DNA samples were polymerase chain reaction amplified and sequenced by the Australian Genome Research Facility. Genotyping was performed as recommended by the manufacturer with reagents included in the iPLEX Gold SNP genotyping kit (Agena) and the software and equipment provided with the MassARRAY platform (Agena) [5]. BDNF serum concentration was analysed using the same methodology as described for analysing serum cytokine levels. Cartridge limits of detection for BDNF serum | ✓ | ✓ | Genotype: Met/Met, Met/Val, Val/Val<br><br>Serum concentration: pg/mL |

| | | | |
|---|---|---|---|
| | concentration were 5.25 pg/ml and samples below this level were allocated a value of zero. | | |
| Serum proteomic profile $^{\Delta}$ | Serum samples for a subgroup of 60 participants with acute LBP were prepared by digesting 3µl of serum (57µg ul-1 +/- 7µg) in 50µl of 50mM AMBIC, 2M urea, 10mM DTT at pH 8 using trypsin at 25°C for 16 hours in a 1:100 enzyme to protein ratio. Serum peptides were fractionated using hydrophobic interaction chromatography (HILIC) according to the manufacturers protocol (PolyLC Inc, MD, USA). Digested and fractionated peptides were reconstituted in 5µL 0.1% formic acid and separated by nano-LC using an Ultimate 3000 HPLC and autosampler (Dionex, Amsterdam, Netherlands). The QExactive (Thermo Electron, Bremen,Germany) mass spectrometer was run in DDA mode. Proteins were identified from the Uniprot database. Protein identifications were accepted if they could be | ✓ | Spectral count, normalised by total ion count |

| | | | |
|---|---|---|---|
| | established at less than 5% FDR and contained at least two identified peptides. | | |
| Genome-wide DNA methylation* Δ | Buchal swabs obtained from the cheek of participants on the day of baseline testing were used to prepare genomic DNA for a subgroup of 60 participants with acute LBP (Isohelix DNA Isolation Kit). Samples were immediately frozen at −80°C and stored. Samples were sent to Australian Genome Research Facility (Melbourne node) where they underwent Quality assessment using QuantiFluor. The samples were then normalised to approximately 250ng of DNA in 45µL and bisulfite converted with Zymo EZ-96 DNA Methylation kit (Zymo Research, Orange, CA). DNA was whole-genome amplified, enzymatically fragmented, purified, and applied to the Illumina MerthylationEPIC BeadChips (Illumina, San Diego, CA) according to the Illumina methylation protocol [3;18]. Beadchips were scanned using the Illumina HiScan | ✓ | Type: Differentially methylated genes |

| | | | | |
|---|---|---|---|---|
| | SQ and the methylation score for each CpG was represented as a β value according to the fluorescent intensity ratio. | | | |
| *Pain processing* | | | | |
| Pressure pain sensitivity $^{\Delta}$ | Laboratory measure: Pressure pain thresholds (PPT) were assessed using a hand help pressure algometer (Somedic, Hörby, Sweden, probe size 1cm$^2$) at three distinct sites: (1) the site of worst LBP (side of most pain on palpation); (2) 3cm lateral to the L3 spinous process on the less painful side of the lower back; and (3) the thumbnail bed (PPT) of the hand contralateral to worst LBP. For pain-free controls, PPTs were measured 3 cm lateral to the L3 spinous process bilaterally and over the thumbnail bed of the dominant hand. Pressure was applied at a rate of 40 kPa/s and participants used a hand-held trigger to indicate when the sensation of pressure first changed to one of pain. Three measures were made at each site and averaged for analysis. | ✓ | ✓ | PPT (kPa, ↑score =↑threshold to pressure pain), |

| | | | | |
|---|---|---|---|---|
| Heat pain sensitivity [Δ] | Laboratory measure: Heat pain thresholds were measured (Thermal Sensory Analyzer, TSA-2001, Q-Sense-CPM, Medoc Ltd, Ramat Yishai, Israel). A 30 x 30 mm Peltier-based thermode was placed on the skin and HPT measured at three site: (1) site of worst LBP, (2) the opposite side of the lumbar region and (3) the ventral aspect of the forearm on the side of worst pain. For pain-free controls, HPTs were measured 3 cm lateral to the L3 spinous process bilaterally and over the ventral aspect of the forearm of the dominant hand. The temperature started at 32°C and increased at a rate of 0.5°C/s. Participants were instructed to push a button when the sensation of heat first changed to one of pain. Three measures were made at each site and the average at each site used for analyses | ✓ | ✓ | HPT (°C, ↑score =↑threshold to heat pain) |

| | | | | |
|---|---|---|---|---|
| Descending pain modulation [Δ] | Laboratory measure: Assessed using an established conditioned pain modulation (CPM) paradigm [13]. PPT was used as the test stimulus (TS) and noxious heat (1°C > HPT) as the conditioning stimulus (CS). Participants completed two trials in random order separated by a 15-min break: (Trial 1) TS at the site of worst LBP and CS on the opposite forearm; (Trial 2) TS at the ipsilateral forearm of worst LBP and CS on the low back opposite to the side of worst pain. In pain-free controls the TS for Trial 1 was the lower back at the level of L3 ipsilateral to the dominant hand and CS on the opposite forearm. For Trial 2, the TS was applied to the forearm of the dominant hand and CS on the low back at the level of L3 opposite the side of TS. Three consecutive PPTs were measured before the application of heat ($TS_1$). Noxious heat was then applied and maintained for the duration of the test, with three consecutive PPTs re-measured 30 seconds | ✓ | ✓ | CPM (kPa, >0 = pain inhibition, <0 = deficient pain inhibition) |

| | | | | |
|---|---|---|---|---|
| | post heat application ($TS_2$). Participants were instructed to rate their pain on a numerical rating scale (0–100) at 0 s, 30 s and immediately following the final PPT measurement. Pain scores were maintained between 50 and 80/100 during testing. The test stimulus was adjusted by 1°C as required to achieve a pain score within this range. The CPM response was calculated as $TS_2$ minus $TS_1$. | | | |
| Nociceptor flexor withdrawal reflex (NFR) [Δ] | Laboratory measure: The NFR was recorded from the biceps femoris muscle on the side of worst LBP (or matched side in pain-free controls). Electrical stimuli were delivered to the sural nerve within the retro-malleolar pathway according to a +/- 20 s variable interval schedule. The NFR threshold was determined as the lowest stimulator intensity that elicited a reflex (4 mA increase until reflex detected, then 2 mA decrease until reflex absent). The stimulus intensity was then set at 120 % of the NFR threshold and five trials recorded. | ✓ | ✓ | Amplitude (mV)<br>Latency (ms) |

| | | | | |
|---|---|---|---|---|
| | The NFR was identified as the multiphasic response occurring 90-200 ms after each stimulus [1;7;21;24] | | | |
| *Lifestyle* | | | | |
| International Physical Activity Questionnaire (IPAQ) [14] $^{\Delta}$ | Questionnaire including seven-items evaluating health-related physical activity. Respondents report the volume of physical activity performed over the previous week, including vigorous activity (activities that make breathing much harder than normal), moderate activity (activities that make breathing somewhat harder than normal), walking and sitting time. | ✓ | ✓ | ↑score = ↑ physical activity (refer to scoring manual for calculating and interpreting MET scores and activity categories) |

* indicates measure was only collected at baseline assessment. All other measures were collected at baseline, 3- and 6-months.

$^{\&}$ indicates measure was collected at baseline and 6-months.

$^{\Delta}$ indicates measure is additional to those reported in the trial registration and study protocol

BDNF – Brain derived neurotrophic factor; CRP – C-reactive protein; EMG – electromyography; IL-1β – interleukin-1 beta; IL-2 – interleukin-2; IL-4 – interleukin-4; IL-6 – interleukin-6; IL-8 – interleukin-8; IL-10 – interleukin-10; 1L-15 – interleukin-15; kPa – kilo Pascal; M1 – primary motor cortex; Met – Methionine; MET – metabolic equivalent of task; TGF-β1 – transforming growth factor beta-1; TNF – tumor necrosis factor; Val – Valine

**Table S2.** Number of baseline and follow-up low back pain participants that provided valid data for all questionnaire items.

| Measure | Baseline (N=120) | 3 months (N=100) | 6 months (N=96) |
|---|---|---|---|
| *Demographic and health* | | | |
| Age (years) | 120 | NA | NA |
| Height (cm) | 111 | NA | NA |
| Weight (kg) | 114 | 73 | 77 |
| Sex | 120 | NA | NA |
| BMI (kg/m$^2$) | 111 | 73 | 76 |
| Comorbidities | 117 | NA | NA |
| Previous LBP | 116 | NA | NA |
| Health care usage | 117 | 95 | 96 |
| Medication usage | 118 | 95 | 96 |
| *Sociodemographic* | | | |
| Cultural diversity | 115 | NA | NA |
| Education | 118 | NA | NA |
| Employment status | 119 | 95 | 93 |
| Impending compensation | 118 | NA | NA |
| Sickness benefits | 110 | 83 | 76 |
| Pain affected work | 117 | 95 | 96 |
| *Pain and disability* | | | |
| Brief pain inventory short form | 118 | 95 | 96 |
| Roland-Morris Disability Questionnaire | 118 | 95 | 96 |
| *Lifestyle* | | | |
| International physical activity questionnaire | 116 | 95 | 96 |

NA indicates questionnaire data was not reassessed at three- and six-month follow-up.
